# Maternal mortality and adverse child outcomes for women with disabilities: A systematic review and meta-analysis

**DOI:** 10.64898/2026.08.25.26361292

**Authors:** Sara Rotenberg, Maureen Chilufya-Moyo, Anne Valentine, Tracey Smythe, Ian Forde, Monika Mitra, Hannah Kuper

**Author notes:** **Declarations of Interest** The authors declare no competing interests. **Funding** Conrad N Hilton Foundation grant (33175), NIHR (NIHR301621), UK Foreign, Commonwealth and Development Office (GB-EDU-133903-PENDA), The National Institute on Disability, Independent Living, and Rehabilitation Research (NIDILRR grant number 90DPHF0013). NIDILRR is a center within the Administration for Community Living (ACL), Department of Health and Human Services (HHS). The content is solely the responsibility of the authors and does not necessarily represent the official views of NIDILRR, ACL, or HHS. **Author Contributions** SR and HK conceptualised the study and wrote the protocol, with input from TS and MMC. SR, HK, and MMC contributed to screening articles. AV and MMC did the data extraction. MMC assessed risk of bias, which was checked by SR. SR conducted the meta-analysis. SR and HK wrote the draft, and all authors meaningfully contributed to the final paper.

## Abstract

**Background:** Reducing maternal mortality and improving newborn and child outcomes are targets of the Sustainable Development Goals. Evidence on how these efforts are reaching women with disabilities is lacking.

**Objectives:** to estimate global, relative inequalities in stillbirth, neonatal, infant, and maternal mortality for women with disabilities compared to women without disabilities.

**Methods:** We searched MEDLINE, Global Health, PsycINFO, and Embase from 1 January 2015 to 28 January 2026, to identify articles on disability and stillbirth, neonatal, infant, and maternal mortality. We included studies that had a recognised measure of disability as an exposure, a control group of women without disabilities and at least one of the four outcomes. A pooled estimate for each outcome was done using a random-effects meta-analysis of the minimally adjusted results.

**Results:** We identified over 4,300 titles, of which 17 papers were eligible for inclusion. Almost all data came from nationally-representative data sources in high-income countries. We found that women with disabilities were 4.66 times more likely (95% C.I. 1.70-12.75) to experience maternal mortality compared to women without disabilities. Women with disabilities were also 37% more likely to have a stillbirth and 27% and 48% more likely to have a neonatal or infant death compared to women without disabilities, respectively.

**Conclusions:** Women with disabilities consistently have higher incidence of maternal, stillbirth, neonatal, and infant mortality, even in countries that have relatively low incidence of these outcomes. There is a lack of evidence globally and particularly from LMICs, and on effective interventions to improve maternal and infant outcomes for women with disabilities.

## Introduction

Reducing maternal and infant mortality is a global priority, embedded within the Sustainable Development Goals (SDGs).^1,2^ An estimated 50% of maternal and infant deaths are avoidable through improving health systems to provide high-quality maternity care.^3^ There is not only a moral imperative to act to reduce the estimated 230,000 maternal^4^ and 2.3 million newborn deaths per year,^5^ but also an opportunity to strengthen societies and economies by enabling women and children to survive, thrive, and build more resilient communities for the future.^6^ Yet, progress is stalling and global targets for these key indicators are unlikely to be reached by 2030.^4^ It is therefore important to identify high risk groups for maternal and infant mortality, to target additional efforts in the spirit of the SDG’s ambition of ‘no one left behind’.

Women with disabilities are one potential high-risk group. This heterogenous group includes women with long-term physical, mental, intellectual, or sensory impairments that, in interaction with societal barriers such as inaccessible environments or negative attitudes, significantly hinder their full participation in society.^7^ This conceptualization of disability helps explain why women with disabilities are likely to experience higher rates of maternal and infant mortality. First, the widespread discrimination and exclusion that women with disabilities face in society means that they frequently have adverse social determinants of health (e.g. lower levels of education, higher levels of poverty), which are known contributors to maternal and infant mortality.^8^ Second, a woman’s impairment or underlying health condition may create physiological risks in pregnancy and childbirth. For instance, women with spinal cord injuries or cerebral palsy, may experience cardiopulmonary or musculoskeletal complications during pregnancy and face substantial barriers to vaginal delivery, increasing the need for caesarean section or other obstetric interventions to ensure safe childbirth..^9^ Third, health systems are rarely built to meet the needs of people with disabilities and health workers are rarely provided with clinical guidance or training on providing disability-inclusive maternal care.^10^ Women with disabilities are likely to face a range of barriers in accessing quality care, such as negative attitudes from health staff on their pregnancy, inaccessible information, equipment and facilities or financial constraints to seeking care.^11^ This third pathway is a result of modifiable health systems barriers that can therefore be addressed through health systems strengthening efforts.

People with disabilities experience higher mortality rates across the life-course and geographies.^12-14^ Poor quality maternal care and higher incidence of complications experienced by disabled women suggest that maternal mortality and adverse childbirth outcomes are another area where these inequalities exist. Previous systematic reviews have also shown that women with disabilities have greater risk of complications from pregnancy than women without disabilities, including gestationaldiabetes and hypertension, and are more likely to require obstetric interventions, such as caesarean sections.^15^ Consistent with these findings, individual studies from high-income settings have shown that women with disabilities are more likely to experience maternal or infant mortality, despite this, the evidence on maternal and infant mortality has not yet been systematically reviewedsynthesised.^16^ For instance, a study in Canada showed that maternal mortality was significantly higher among women with disabilities compared to those without (minimally adjusted RR 1.77, 95% CI: 1.17-2.69).^17^ Similarly, in the USA, evidence shows that maternal mortality rates are higher for women with disabilities, ^18^ as well as for women with specific disability types, such as intellectual and developmental disabilities^19^ and physical disabilities.^20^ The data on infant mortality appears relatively limited,^15^ but there is emerging evidence that women with psychosocial disabilities are more likely to experience stillbirth or infant mortality.^21^

Although these studies provide important evidence on maternal health inequalities experienced by women with disabilities, data from low- and middle-income countries remain strikingly limited, despite the fact that 90% of maternal deaths occur in these settings^4^. It is particularly concerning that 80% of people with disabilities live in LMICs —where the burden of maternal mortality is greatest - yet where evidence on disability is most sparse. Consequently, the aim of this study is to undertake a global systematic review and meta-analysis to synthesise the evidence on the association between disability status and adverse perinatal outcomes, including stillbirth and maternal, neonatal, and infant mortality.

## Methods

We undertook a systematic review and meta-analysis to compare the incidence of four outcomes: stillbirth and maternal, neonatal, and infant mortality in women with disabilities compared with women without disabilities or women in the general population. We conducted a meta-analysis on all four outcomes. We followed PRISMA reporting guidelines and registered the systematic review and meta-analysis with PROSPERO (CRD420261329635).

### Definitions

Disability was defined using the United Nations Convention on the Rights of Persons with Disabilities, which defines people with disabilities as “those who have long-term physical, mental, intellectual or sensory impairments which in interaction with various barriers may hinder their full and effective participation in society on an equal basis with others.”^7^ This definition includes people with specific conditions deemed likely to result in disability (e.g., spina bifida, muscular dystrophy, Down Syndrome), specific impairments (e.g. visual, hearing, physical) as well as disability measured through functioning/activity limitations/self-report (e.g., Washington Group questions, activities of daily living). We excluded papers focussed on people with mild functional impairments, frailty, or specific diseases that have a range of functioning (e.g., Parkinson’s, anxiety, depression, etc.)

We defined maternal mortality according to the WHO International Classification of Diseases (ICD-10) as the death of a woman during pregnancy and childbirth or within 42 days of termination of pregnancy, from any cause related to or aggravated by pregnancy or its management (excluding accidental or incidental causes), irrespective of the duration and site of the pregnancy.^22^ Infant mortality was defined as death of a live-born infant between birth and 11 months.^23^ Neonatal mortality was defined as death among live-born infants during the first 28 days of life.^24^ We defined stillbirth in accordance with international reporting standards as a baby born with no signs of life at 28 weeks or more of gestation.^25^

### Inclusion and exclusion criteria

We included published prospective and retrospective cohort studies, cross-sectional studies, and randomised controlled trials with a baseline assessment of disability in women and a longitudinal assessment of maternal and/or infant mortality. The eligible effect measures for the association between disability and maternal/infant mortality included hazard ratio (HR), cumulative incidence ratio (CIR) or odds ratio (OR) with 95% CIs, or standardised mortality ratios. We excluded studies that did not include a referent group of nondisabled women or women in the general population. There were no restrictions in language of publication or country or setting (e.g. community based versus hospital based). There were no restrictions on the age of the woman, but most studies would focus on those in the reproductive age group (15-49). Studies that were excluded and their reasons for exclusion are in Appendix 4.

### Search strategy, selection criteria and data extraction

We searched MEDLINE, Global Health, PsycINFO, and Embase from 1 January 2015 until 28 January 2026, to identify articles on disability and the four outcomes. The start date of 1 January 2015 was selected as it marks the start of the 2030 Agenda for Sustainable Development to reach the Sustainable Development Goals, which represents a renewed focus on reducing maternal and infant mortality. Results before this date were therefore deemed to be outdated within these efforts. Search terms were piloted in the Ovid search platform and can be found in Appendix 1. The search process and study protocol were registered on PROSPERO on 9 March 2026 prior to extraction finishing. The study selection was conducted by two researchers (SR and HK). All titles and abstracts were systematically screened against the eligibility criteria in Covidence. Next, two reviewers (SR and MMC) independently assessed the full text of each article for inclusion.

Three researchers (SR, AV, and MMC) developed and pilot-tested an extraction tool in Excel to systematically record information from the included studies. Extracted information included: 1) publication characteristics (author, title, year of publication, country or location, study name); 2) study design (study design, sample size, source of data, follow-up duration); 3) participant characteristics (age, disability measurement); and 4) outcomes. For each outcome, we extracted the numerator (number of maternal, infant, neonatal deaths, or stillbirths) and the denominator used to calculate the effect estimate (e.g., number of live births, pregnancies), together with the postpartum time window and method of outcome ascertainment. Table 2 summarises the extraction table, while the full extraction table is in Appendix 2.

**Table 1:**
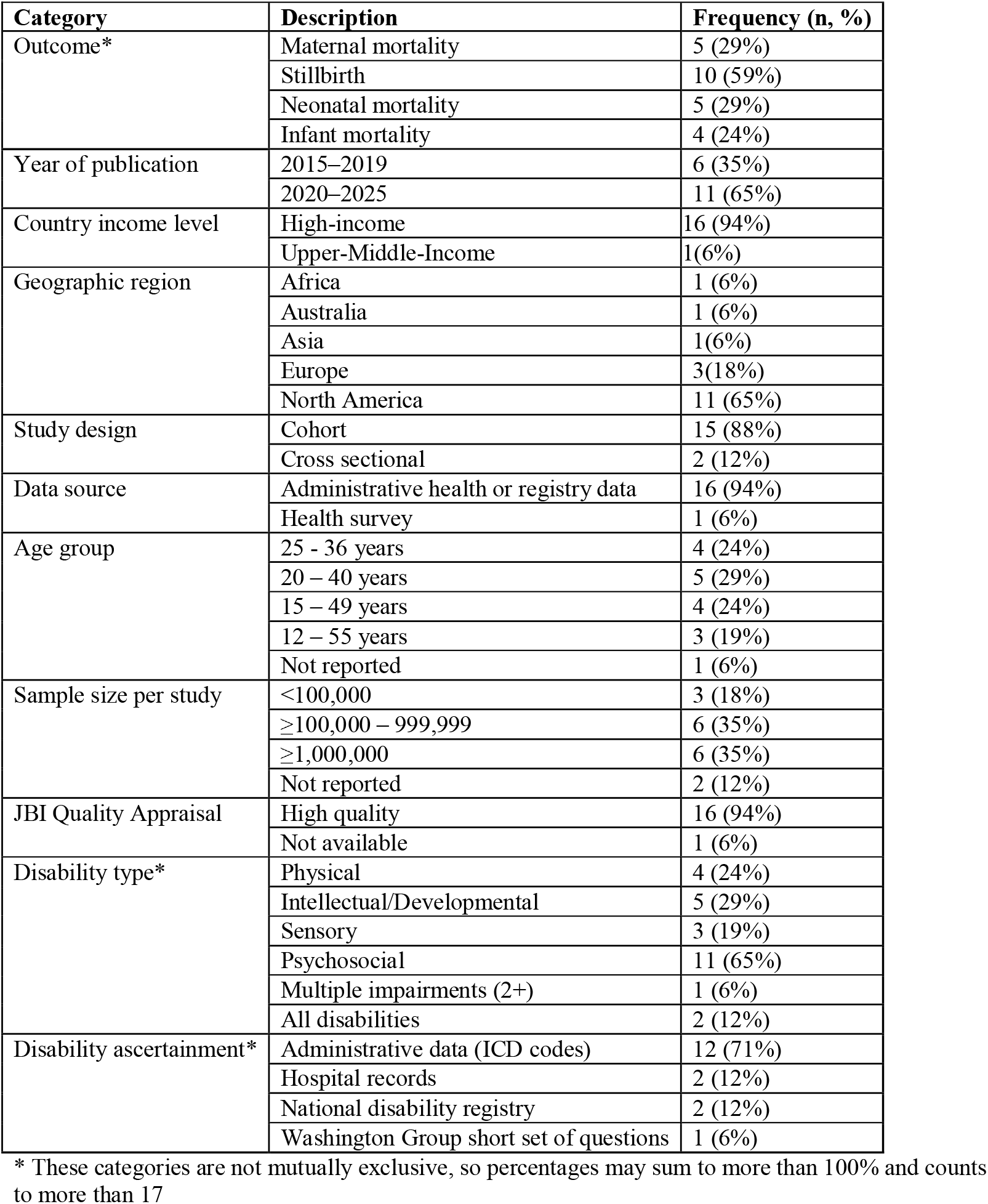
Characteristics of Included Studies (N=17)

**Table 2:**
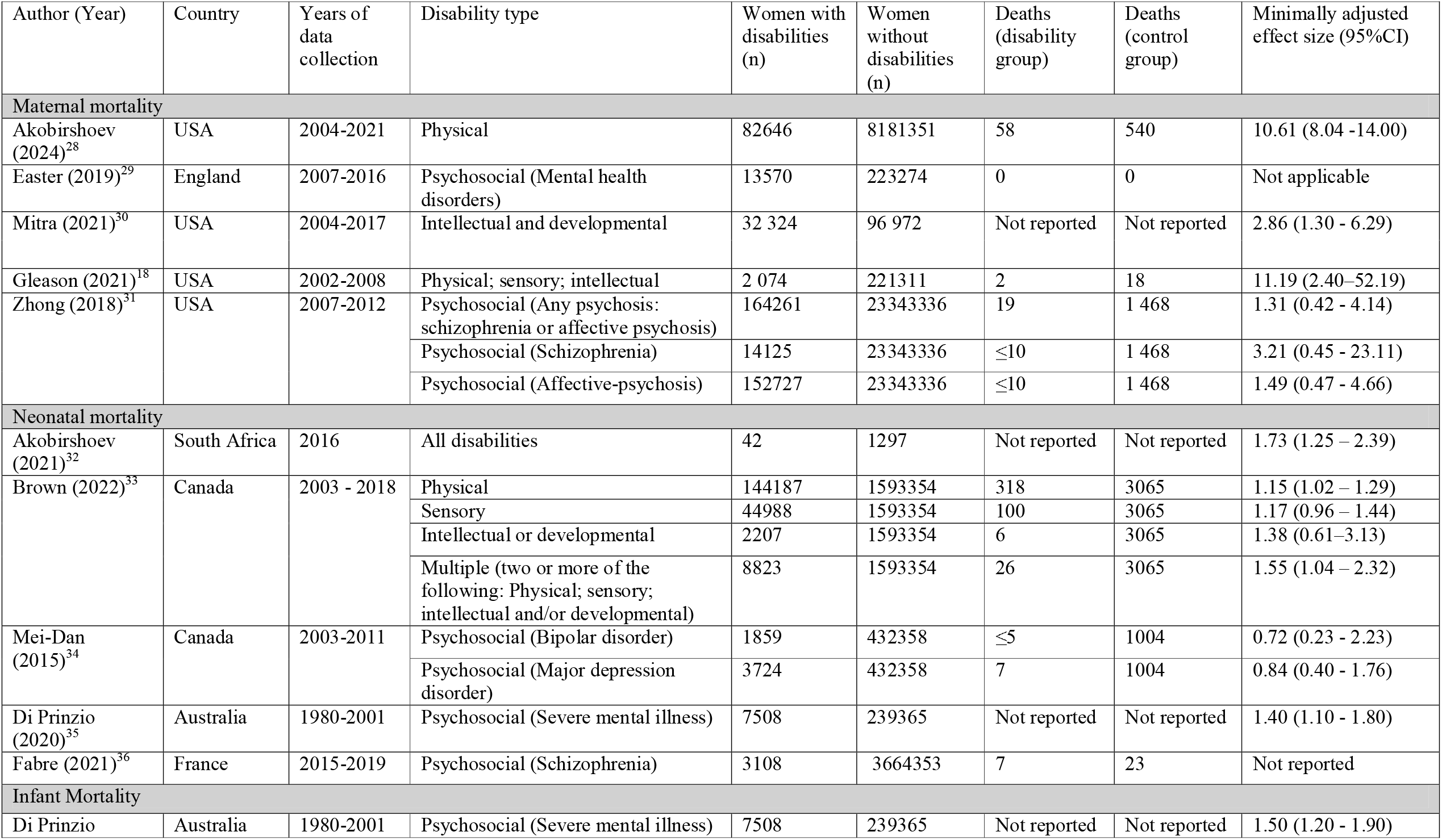
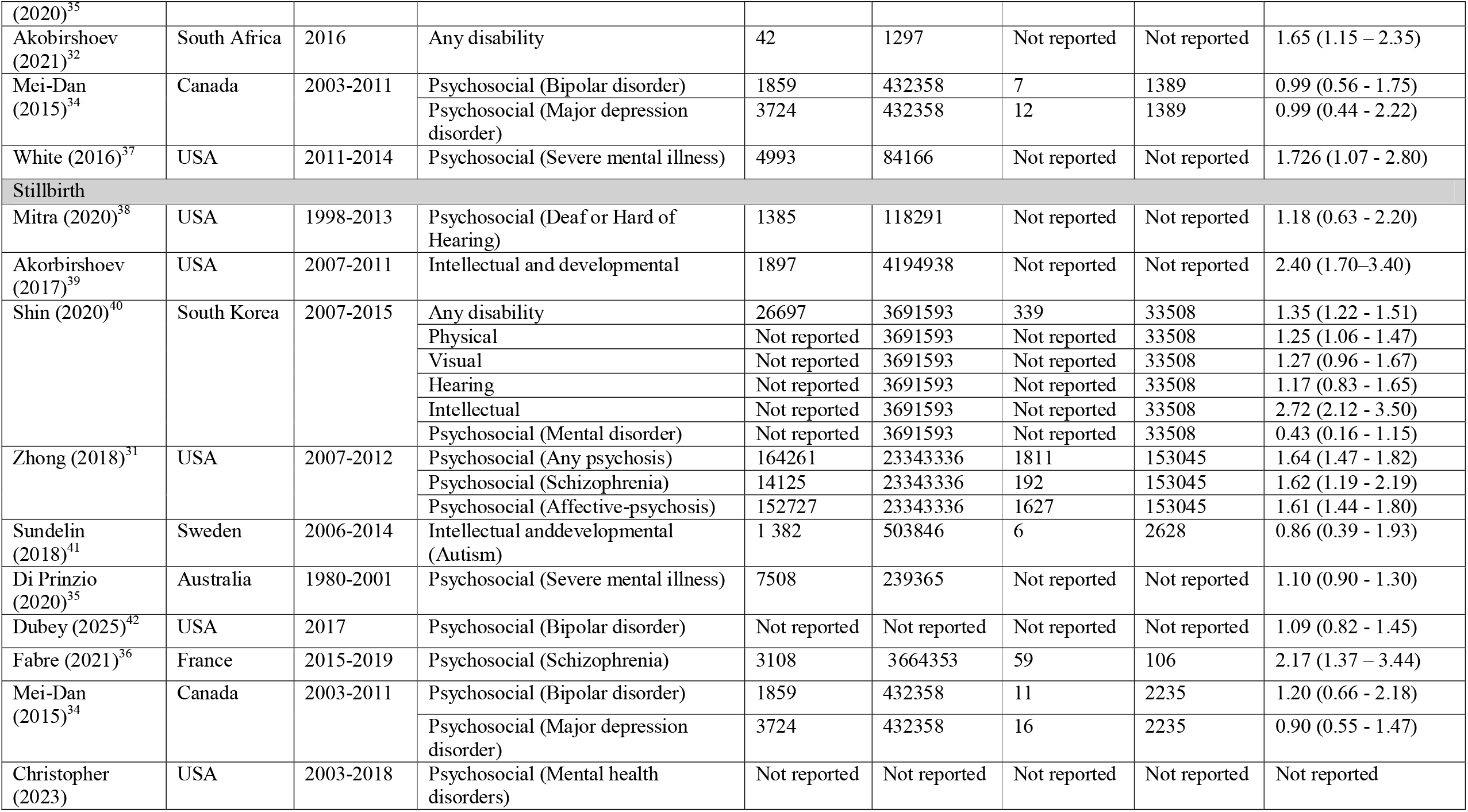
Summary of studies included in meta-analysis.

We extracted all reported effect estimates (number, relative risk [eg, HR, CIR, OR] with corresponding 95% confidence intervals, and we extracted estimates with different levels of adjustment [eg, age -adjusted and multivariate adjusted]). We selected the least adjusted estimate for the multivariate models if several models were reported, given the heterogenous covariates available and used in each model. Where effect estimates were reported without explicit numerators or denominators, we extracted the reported relative measure and recorded the underlying denominator where stated. Data were extracted for each included study independently by one reviewer (AV) and checked by a second reviewer (MMC), with discrepancies discussed and resolved.

### Risk of Bias

One researcher (MMC)graded the overall certainty of the evidence using the relevant Joanna Briggs Institute Critical Appraisal Checklist (either cohort or cross-sectional)^26,27^, which was checked by a second researcher (SR) (Appendix 3). The requirement to demonstrate that the outcome of interest was absent at baseline was waived, as it was mortality. We assigned a score of low, medium, or high risk of bias for each criterion, with the overall score assigned on the basis of the worst score given for any individual item; a score of high was given if there were at least two medium scores or at least one high score.

### Meta-analysis

We undertook a random-effects meta-analyses to generate a pooled effect estimate, with 95% CIs for the association between maternal disability and the four outcomes of maternal mortality, stillbirth, neonatal mortality, and infant mortality, if there were at least three studies for each outcome. Meta-analyses were undertaken only when outcome definitions, postpartum time windows, and denominators were sufficiently comparable. Effect measures differed across studies (for example, relative risk versus odds ratios), but they were treated as equivalent on the advice of a statistician given the binary nature of the outcome as mortality and the similar outcome and exposure definitions. Between-study heterogeneity was assessed with *I*^2^ and τ^2^ statistics. We conducted the meta-analyses using the minimally adjusted estimate (at least age and often socio-economic status), as that was most comparable and available for all but one study, where we used the unadjusted estimate. We used the overall estimate of disability in each study when it was available, or the impairment-disaggregated ones where only that was reported. We planned to undertake sub-group meta-analyses by type of disability, if there were at least 5 studies identified that reported on the results, but this was not possible with the results available. We assessed the risk of publication bias by inspection of funnel plots. We did all statistical analyses using R Version 4.4.1.

### Role of the funding source

The funders of the study had no role in study design, data collection, data analysis, data interpretation, or writing of the report.

## Results

We identified 4,388 studies published between January 2015 and January 2026, and 3,309 were screened once duplicates were removed. After title and abstract screening, 55 full-text studies were assessed for eligibility and 35 were excluded, because they did not examine the outcomes of interest (n=15) or did not include a measure of disability (n=9). Three full texts were ultimately excluded at the extraction stage because they did not compare women with and without disabilities, leaving 17 studies eligible for inclusion in the review, including many with multiple outcomes of interest.

Stillbirth was included in 10 studies, maternal and neonatal mortality in 5 studies, respectively, and infant mortality in 4 studies. Nine studies were published from 2015-2020, while eight studies were published between 2021 and 2026 . Most studies were conducted in high income countries (94%) and mostly from North America (65%), though there was one study from an upper-middle income country (South Africa). The majority of studies (88%) were cohort studies utilizing administrative data or health data sources;, and there wasone study from a National Demographic and Health Survey. Included studies were large, with most over 100,000 people and several over one million women. Disability was predominantly ascertained via ICD codes in administrative data or other hospital or disability records, and psychosocial disability was the most common type of impairment (65%) included in the exposure group, with several studies also examining intellectual or developmental disability (29%). All studies were rated as high-quality using the relevant JBI quality appraisal checklist, except for one conference abstract that could not be evaluated.

Our meta-analysis (Figure 2) shows women with disabilities were more likely to experience all four adverse pregnancy-related outcomes compared to women without disabilities. One maternal mortality study did not have any deaths in either group, but the remaining three showed statistically significant elevated mortality rate for women with disabilities. Overall, women with disabilities had 4.66 (95% C.I. 1.70-12.75) times higher maternal mortality rate compared to women without disabilities. Moreover, across 10 studies and 11 cohorts, women with disabilities had 1.37 (95% C.I. 1.17-1.61) times higher incidence of stillbirth compared to women without disabilities. IInfant mortality was reported in four studies for 5 cohorts, showing that women with disabilities had 48% higher incidence of death of a live-born child under one year (RR: 1.48, 95% C.I. 1.25-1.75) compared to women without disabilities. Finally, neonatal mortality was reported for nine cohorts within five studies. Women with disabilities had a 27% higher incidence of neonatal mortality (RR: 1.27, 95% C.I. 1.12-1.44) compared to women without disabilities. Between-study heterogeneity varied across meta-analyses and was highest for maternal mortality (I^2^=85%) and lowest for infant mortality (I^2^=0%). Funnel plots for each of the four outcomes showed no evidence of publication bias (Appendices 6-8)

**Figure 1:**
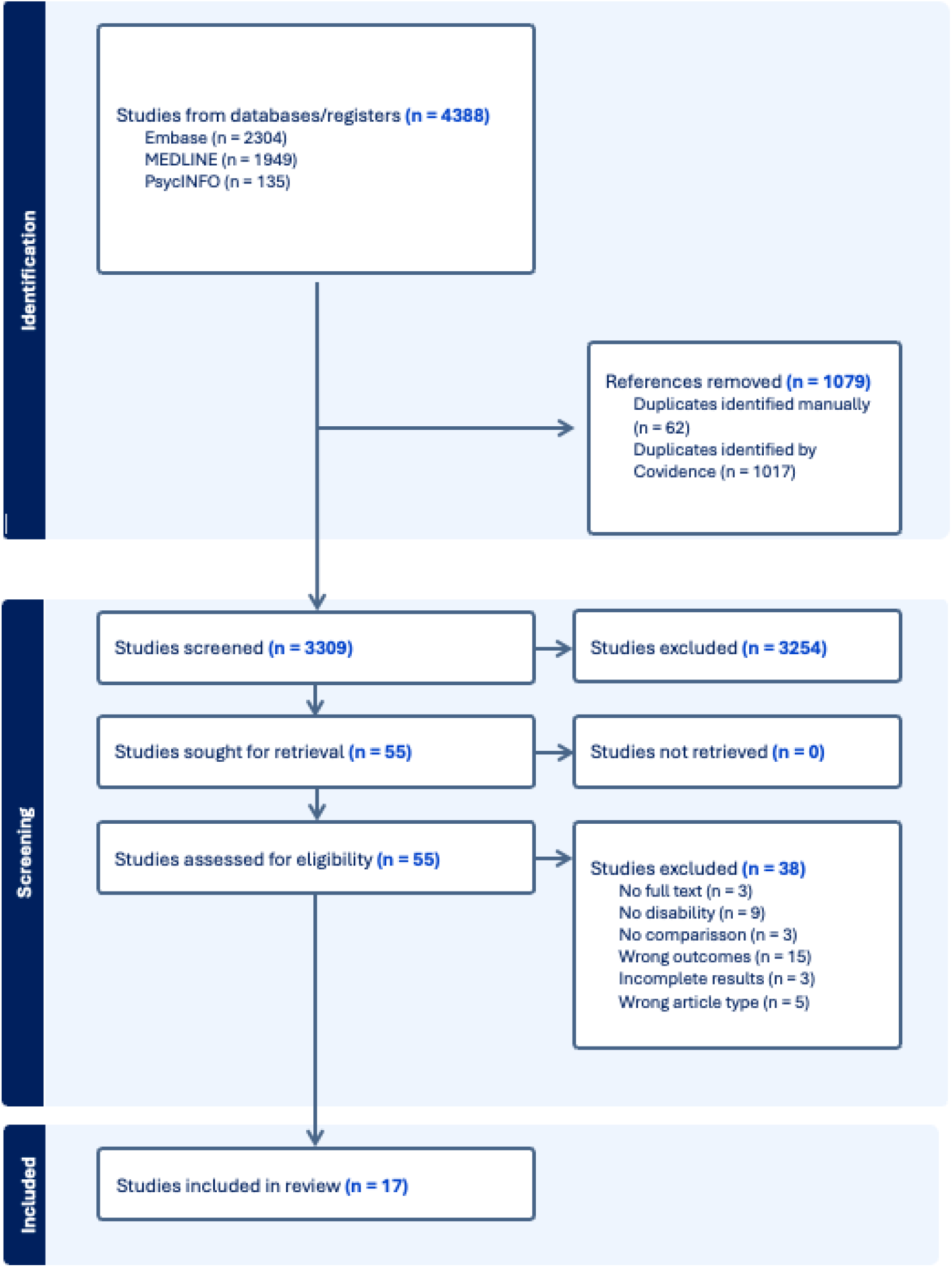
PRISMA Flowchart.

**Figure 2:**
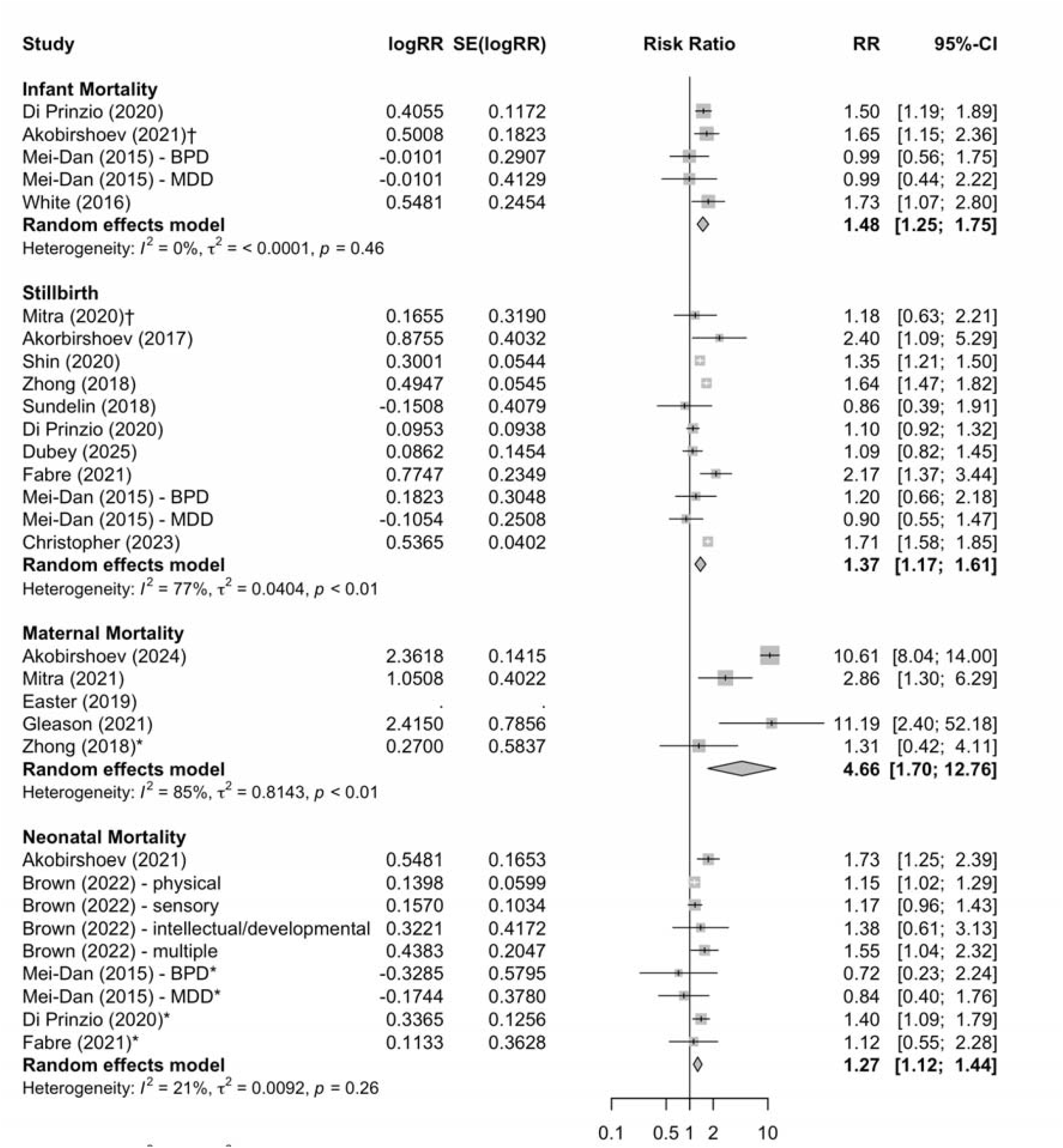
Meta-analysis results by outcome.

## Discussion

Women with disabilities have equal sexual and reproductive health rights as those without disabilities, including the right to safe, respectful and accessible childbirth and newborn and infant care. However, the results this systematic review and meta-analysis highlight persistent inequalities in reproductive care for women with disabilities. Our results show that women with disabilities from high-income countries are significantly more likely to experience maternal mortality, stillbirth, and neonatal and infant mortality. These outcomes are rare and individual studies often show higher levels for women with disabilities, but not significant differences, highlighting the importance of this synthesised, meta-analysis. Previous reviews have demonstrated the increased risk of pregnancy and postpartum complications, including gestational diabetes, hypertension, and obstetric intervention, such as caesarean delivery.^15^ These results provide newly synthesised evidence on mortality outcomes that reflect the broader inequities in access to and quality of maternal care for women with disabilities, as well as the cumulative effects of lifelong structural disadvantage for women with disbailities.^43^

These findings show that relative inequalities for women with disabilities are starkest for maternal mortality, where women are nearly five times more likely to experience maternal mortality compared to women without disabilities. Although study heterogeneity was high for this outcome, the directionally uniform and statistically significant elevation risk across all studies is important. Future studies with consistent disability ascertainment methods and adjustment strategies will be needed to produce a more stable pooled estimate of the hightened risk of maternal mortality. Indeed, there were only five studies identified on this important topic, all of which came from either the United States^18,28,30,31^ or United Kingdom,^29^ making two salient points about data availability and geography. First, these unacceptable inequities persist for women with disabilities even in countries with relatively low incidence of these adverse maternal outcomes.^4^. The persistence of these inequities highlights that reductions in maternal mortality at the population level have not been equitably experienced by women with disabilities, emphasising the need for targeted and disability-inclusive interventions. Second, 90% of maternal mortality occurs in low- and middle-income countries^4^, where 80% of people with disabilities live, yet there was not a single study on maternal mortality from the global South and only five studies on this topic overall. The lack of data on this critical public health outcome represents a critical gap, especially considering there is ample evidence of inequities in maternal health care for women with disabilities in low- and middle-income settings.^44,45^ More data are needed globally to understand and address this issue, particularly well-powered studies that allow exploration of intersectional factors, like poverty and ethnicity, as has been done in the USA.^20,46^ Without population-level data, it is difficult to ascertain whether improvements in maternal care are systemically reaching women with disabilities in low- and middle-income countries.

Moreover, our results demonstrate 30-50% greater risk of stillbirth, neonatal mortality, and infant mortality among births to women with disabilities compared to women without disabilities. The reasons behind these disparities are likely complex, while the health conditions associated with some disabilities may contribute to increased physiological risk in pregnancy, the evidence for this remains limited.^18^ However, the evidence of inequities across disability type suggests this pattern is not merely a biological phenomenon, but rather a confluence of complex social factors. The social determinants that drive these outcomes reflect the stigma, discrimination and structural barriers that disabled women experience across the life course. For example, prior reviews have shown that stillbirth is associated with poverty, lower education, HIV status, and diabetes.^47^ Women with disabilities are more likely to live in poverty,^48^ have lower educational attainment, and have higher prevalence of co-morbidities, including diabetes and HIV,^49-51^ these inequities help to explain the elevated risk for adverse outcomes observed in this review. However, given that we used results that adjusted for some of these factors, it suggests that some of this inequity is because of how health systems are unprepared to serve women with disabilities. This includes inaccessible facilities and equipment, limited provider training in disability-inclusive care, and an absence of practice guidelines for providers. Multiple studies have shown that women with disabilities would benefit from greater conception, pregnancy, and post-natal support, but are unable to access it.^17,52-54^ Therefore, greater attention to providing high-quality, targeted maternal care for disabled women is essential to reducing these largely preventable inequalities.

Despite this evidence on inequalities, a recent systematic review found no eligible intervention studies designed to address disability-related barriers during pregnancy and childbirth.^55^ General health systems strengthening interventions for people with disabilities, such as health worker training to improve attitudes, improving accessibility of health facilities, accessible transport, and financial protection, may support imrpovements to maternity care for women with disabilities.^51^ However, it is clear lack of data^56^ are at the crux of both the problem and the solutions. Therefore, there is an urgent need to not only invest in vital registration systems that collect disability data to understand maternal, neonatal, and infant mortality and stillbirth by maternal disability status, but also to invest in research to develop targeted interventions and more informed policy guidance for health practitioners.^10^ In high-income settings, priority actions should include mandating disability assessment or screening at antenatal booking; linking national disability registers and birth/hospital episode data; and adding disability as a mandatory field in confidential maternal death enquiries. Countries currently developing their civil registration and vital statistics measures have a prime opportunity to integrate disability data collection into the design and implementation of these programmes, to fill this critical gap in evidence. In low and middle-income settings, integrating standardised question sets, such as Washington Group questions into household survey birth history modules; embedding disability-disaggregated maternal and child health indicators into health management information and civil vital registration reform programmes will create meaningful opportunities to improve these findings.

### Strengths and Limitations

This review used a tested search strategy and reputable data sources to pool recent evidence on important maternal indicators for women with disabilities compared to women without disabilities. By limiting our inclusion criteria to only studies with a referent group, we provide evidence on the relative inequalities for women with disabilities compared to women without disabilities. However, this study has notable limitations. First, we included only one study from an upper-middle income country, limiting the global applicability of our findings. Second, many included studies reported estimates for disability overall, limiting our ability to identify which women with disabilities experience the greatest risk. Only a handful of studies disaggregated by disability type and current evidence is skewed toward psychosocial disabilities (65% of studies) and lacks sufficient detail on sensory or physical impairments. Data were minimally adjusted and not disaggregated by other social factors (e.g., race, ethnicity, health status or socio-economic status), which may be important for understanding intersectional inequalities and whether predominantly socio-economic or health service determinants are driving the increased RRs observed. Future research should address the gap in global, disability-specific, and intersectional research.

## Conclusion

Stillbirth, maternal, neonatal, and infant mortality serve as an important barometer for the access and quality of healthcare for women. Across 17 studies, we have demonstrated that women with disabilities have significantly greater risk of these outcomes compared to women without disabilities. While the findings are limited to high-income countries, the absence of data should not be an excuse for inaction, but rather an urgent call to action to this under reported global health issue, and inspire targeted investments in data collection and better maternal care support for women with disabilities globally.

## Data Availability

All data produced in the present study are available upon reasonable request to the authors

## Data statement

The extraction sheet is included in the Supplementary Files. All analysis code are available upon request.

## AI Statement

No AI was used in writing the manuscript.

## Notes

### Competing Interest Statement

The authors have declared no competing interest.

