## Appendix 1 for "Maternal mortality and adverse child outcomes for women with disabilities: A systematic review and meta-analysis"

Appendix 1: Sample Search Strategy

Appendix 2: Full Extraction Table

Appendix 3: Joanna Briggs Institute Quality Appraisal Checklist

Appendix 4: List of Excluded Studies

Appendix 5: Funnel Plot for Maternal Mortality Meta-analysis

Appendix 6: Funnel Plot for Neonatal Mortality Meta-analysis

Appendix 7: Funnel Plot for Infant Mortality Meta-analysis

Appendix 8: Funnel Plot for Stillbirth Meta-analysis

Appendix 1: Sample Search Strategy

| Searches | Results | Type |
| --- | --- | --- |
| 1 | (maternal adj2 (death* or mortalit*)).ti,ab. | 21837 |
| 2 | exp Maternal Mortality/ | 12120 |
| 3 | exp Infant Mortality/ | 32942 |
| 4 | exp neonatal mortality/ | 32942 |
| 5 | infant mortalit*.ti,ab. | 12470 |
| 6 | (neonatal mortalit* or perinatal mortalit* or early neonatal death* or late neonatal death*).ti,ab. | 21456 |
| 7 | (stillbirth* or still-birth* or intrauterine fetal death* or intra-uterine fetal death* or IUFD or fetal death*).ti,ab. | 26397 |
| 8 | exp Disabled Persons/ | 79672 |
| 9 | (disabilit* or handicap* or impair*).ti,ab. | 1286855 |
| 10 | (physical disabilit* or mobility impair* or wheelchair* or musculoskeletal disorder*).ti,ab. | 32486 |
| 11 | (sensory disabilit* or hearing impair* or deaf* or visual impair* or blind*).ti,ab. | 462073 |
| 12 | (intellectual disabilit* or learning disabilit* or developmental disabilit* or cognitive impair*).ti,ab. | 160391 |
| 13 | (autism spectrum disorder* or autis*).ti,ab. | 81185 |
| 14 | (mental illness or serious mental illness or severe mental illness or psychotic disorder* or schizophrenia or bipolar).ti,ab. | 246768 |
| 15 | (psychiatric disabilit* or psychosocial disabilit*).ti,ab. | 1385 |
| 16 | (multiple disabilit* or complex disabilit*).ti,ab. | 1155 |
| 17 | ((maternal or mother* or pregnan* or perinatal or antenatal or women* or woman*) adj3 (disabilit* or impair* or handicap* or chronic condition* or long-term condition*)).ti,ab. | 9443 |
| 18 | 8 or 9 or 10 or 11 or 12 or 13 or 14 or 15 or 16 or 17 | 1993784 |
| 19 | 1 or 2 or 3 or 4 or 5 or 6 or 7 | 92923 |
| 20 | 18 and 19 | 4049 |
| 21 | limit 20 to yr="2015 -Current" | 1947 |

Appendix 2: Full Extraction Table

| Author (Year) | Country | Study design | Years of data collection | Disability measure | Data source | Disability type | Maternal age (years) | Women with disabilities (n) | Women without disabilities (n) | Deaths (disability) | Deaths (control) | Effect size measure | Unadjusted effect size (95%CI) | Minimally adjusted effect size (95%CI) | Fully adjusted effect size (95% CI) |
| --- | --- | --- | --- | --- | --- | --- | --- | --- | --- | --- | --- | --- | --- | --- | --- |
| **Maternal mortality** | | | | | | | | | | | | | | | |
| Akobirshoev (2024) | USA | Retrospective Cohort | 2004-2021 | ICD-9, ICD 10 codes | National hospital database | Physical | <20 - 40+ | 82646 | 8181351 | 58 | 540 | Relative Risk | Not reported | 10.61 (8.04 -14.00) | 7.75 (5.72-10.50) |
| Easter (2019) | England | Retrospective Cohort | 2007-2016 | ICD 10 codes | National hospital database | Mental health disorders | 22.5 - 36.6 | 13570 | 223274 | 0 | 0 | Not applicable | Not applicable | Not applicable | Not applicable |
| Mitra (2021) | USA | Retrospective Cohort | 2004-2017 | ICD-9, ICD 10 codes | National hospital database | Intellectual and developmental | <20 - 40+ | 32 324 | 96 972 | Not reported | Not reported | Risk Ratio | Not reported | 2.86 (1.30 - 6.29) |  |
|  |  |  |  |  |  |  |  |  |  |  |  |  |  |  | 2.30 (1.05 - 5.29) |
| Gleason (2021) | USA | Retrospective Cohort | 2002-2008 | ICD-9 codes | National hospital database | Physical; sensory; intellectual | Mean (SD): 27.6 (6.2) | 2 074 | 221311 | 2 | 18 | Relative Risk | Not reported | 11.19 (2.40–52.19) | Not reported |
| Zhong (2018) | USA | Retrospective Cohort | 2007-2012 | ICD-9 codes | National hospital database | Any psychosis (schizophrenia or affective psychosis) | 12 - 55 | 164261 | 23343336 | 19 | 1 468 | OR | 1.82 (0.67 - 4.92) | 1.31 (0.42 - 4.14) | 1.00 (0.30 - 3.31) |
|  |  |  |  |  |  | Schizophrenia |  | 14125 | 23343336 | ≤10 | 1 468 | OR | 5.15 (0.73 - 36.61) | 3.21 (0.45 - 23.11) | 2.23 (0.30 - 16.80) |
|  |  |  |  |  |  | Affective-psychosis |  | 152727 | 23343336 | ≤10 | 1 468 | OR | 1.96 (0.72 - 5.29) | 1.49 (0.47 - 4.66) | 1.13 (0.34 - 3.74) |
| **Neonatal mortality** | | | | | | | | | | | | | | | |
| Akobirshoev (2021) | South Africa | Cross sectional | 2016 | Washington Group Short Set of questions (a lot of difficulty/ cannot function at all) | National Demographic and Health Surveys | All disabilities | 15 - 49 | 42 | 1297 | Not reported | Not reported | Risk Ratio | 1.80 (1.31 – 2.49) | 1.73 (1.25 – 2.39) | Not reported |
| Brown (2022) | Canada | Cohort | 2003 - 2018 | Medical records | Linked health adminstrative and hospital dataset | Physical | 15 - 49 | 144187 | 1593354 | 318 | 3065 | Risk Ratio | 1.14 (1.02 – 1.28) | 1.15 (1.02 – 1.29) | 1.12 (0.99 – 1.26) |
|  |  |  |  |  |  | Sensory |  | 44988 | 1593354 | 100 | 3065 | Risk Ratio | 1.15 (0.94 – 1.41) | 1.17 (0.96 – 1.44) | 1.15 (0.94 – 1.41) |
|  |  |  |  |  |  | Intellectual or developmental |  | 2207 | 1593354 | 6 | 3065 | Risk Ratio | 1.37 (0.60–3.10) | 1.38 (0.61–3.13) | 1.27 (0.56 – 2.89) |
|  |  |  |  |  |  | Multiple (two or more of the following: Physical; sensory; intellectual and/or developmenta) |  | 8823 | 1593354 | 26 | 3065 | Risk Ratio | 1.53 (1.03 – 2.29) | 1.55 (1.04 – 2.32) | 1.46 (0.98 – 2.18) |
| Mei-Dan (2015) | Canada | Cohort | 2003-2011 | ICD-9, ICD-10 codes | Provincial health dataset | Bipolar disorder | 14 - 50 | 1859 | 432358 | ≤5 | 1004 | OR | Not reported | 0.72 (0.23 - 2.23) | Not reported |
|  |  |  |  |  |  | Major depression disorder | 14 - 50 | 3724 | 432358 | 7 | 1004 | OR | Not reported | 0.84 (0.40 - 1.76) | Not reported |
| Di Prinzio (2020) | Austrailia | Cohort | 1980-2001 | ICD-9 codes | State hospital registry and mental health information system | Severe mental illness | <20 - 35+ | 7508 | 239365 | Not reported | Not reported | OR | 1.70 (1.30 - 2.10) | 1.40 (1.10 - 1.80) | 1.30 (1.10 - 1.70) |
| Fabre (2021) | France | Matched cohort | 2015-2019 | ICD-10 codes | National hospital discharge data | Schizophrenia | 15 - 35+ | 3108 | 3664353 | 7 | 23 | OR | 1.12 (0.55 – 2.28) | Not reported | Not reported |
| **Infant Mortality** | | | | | | | | | | | | | | | |
| Di Prinzio (2020) | Austrailia | Cohort | 1980-2001 | ICD-9 codes | State hospital registry and mental health information system | Severe mental illness | <20 - 35+ | 7508 | 239365 | Not reported | Not reported | OR | 2.10 (1.70 - 2.70) | 1.50 (1.20 - 1.90) | 1.40 ( 1.10 - 1.80) |
| Akobirshoev (2021) | South Africa | Cross sectional | 2016 | Washington Group Short Set of questions (a lot of difficulty/cannot function at all) | National Demographic and Health Surveys | All disabilities | 15 - 49 | 42 | 1297 | Not reported | Not reported | Risk Ratio | 1.69 (1.19 – 2.41) | 1.65 (1.15 – 2.35) | Not reported |
| Mei-Dan (2015) | Canada | Cohort | 2003-2011 | ICD-9, ICD-10 codes | Provincial health dataset | Bipolar disorder | 14 - 50 | 1859 | 432358 | 7 | 1389 | OR | Not reported | 0.99 (0.56 - 1.75) | Not reported |
|  |  |  |  |  |  | Major depression disorder | 14 - 50 | 3724 | 432358 | 12 | 1389 | OR | Not reported | 0.99 (0.44 - 2.22) | Not reported |
| White (2016) | USA | Cohort | 2011-2014 | Claim form examination | Medical claims data | Severe mental illness | Not reported | 4993 | 84166 | Not reported | Not reported | OR | Not reported | 1.726 ( 1.07 - 2.80) | Not reported |
| **Stillbirth** | | | | | | | | | | | | | | | |
| Mitra (2020) | USA | Cohort | 1998-2013 | ICD-9 codes | Regional longitudinal database | Deaf or Hard of Hearing | <20 - 40+ | 1385 | 118291 | Not reported | Not reported | Risk Ratio | 1.35 (0.67 - 2.69) | 1.18 (0.63 - 2.20) | 1.30 (0.70 - 2.43) |
| Akorbirshoev (2017) | USA | Cohort | 2007-2011 | ICD-9 codes | National health database | Intellectual and developmental | <25 - 34 | 1897 | 4194938 | Not reported | Not reported | OR | 3.52 (2.61 - 4.74) | 2.40 (1.70–3.40) | Not reported |
| Shin (2020) | South Korea | Cohort | 2007-2015 | National Disability Registry | Merged National dataset | All disabilities | 27 - 36 | 26697 | 3691593 | 339 | 33508 | OR | 1.41 (1.26-1.57) | 1.35 (1.22 - 1.51) | 1.30 (1.17–1.45) |
|  |  |  |  |  |  | Physical | 27 - 36 | Not reported | 3691593 | Not reported | 33508 | OR | 1.32 (1.13 - 1.56) | 1.25 (1.06 - 1.47) | 1.22 (1.04 - 1.43) |
|  |  |  |  |  |  | Visual | 27 - 36 | Not reported | 3691593 | Not reported | 33508 | OR | 1.31(1.00 - 1.72) | 1.27 (0.96 - 1.67) | 1.23 (0.94 - 1.62) |
|  |  |  |  |  |  | Hearing | 27 - 36 | Not reported | 3691593 | Not reported | 33508 | OR | 1.21 (0.86 - 1.70) | 1.17 (0.83 - 1.65) | 1.16 (0.83 - 1.64) |
|  |  |  |  |  |  | Intellectual | 27 - 36 | Not reported | 3691593 | Not reported | 33508 | OR | 2.59 (2.01 - 3.33) | 2.72 (2.12 - 3.50) | 2.64 (2.05 - 3.40) |
|  |  |  |  |  |  | Mental disorder | 27 - 36 | Not reported | 3691593 | Not reported | 33508 | OR | 0.47(0.18 - 1.26) | 0.43 (0.16 - 1.15) | 0.41 (0.15 - 1.08) |
| Zhong (2018) | USA | Retrospective Cohort | 2007-2012 | ICD-9 codes | National hospital database | Any psychosis | Dec-55 | 164261 | 23343336 | 1811 | 153045 | OR | 1.69 (1.52 - 1.88) | 1.64 (1.47 - 1.82) | 1.37 (1.23 - 1.53) |
|  |  |  |  |  |  | Schizophrenia | Dec-55 | 14125 | 23343336 | 192 | 153045 | OR | 2.09 (1.54 - 2.83) | 1.62 (1.19 - 2.19) | 1.28 ( 0.94 - 1.74) |
|  |  |  |  |  |  | Affective-psychosis | Dec-55 | 152727 | 23343336 | 1627 | 153045 | OR | 1.63 (1.46 - 1.82) | 1.61 (1.44 - 1.80) | 1.35 (1.20 - 1.51) |
| Sundelin (2018) | Sweden | Cohort | 2006-2014 | National Patient Registry | National Birth & Patient Registry | Autism | ≤24 - 35+ | 1 382 | 503846 | 6 | 2628 | OR | 0.91 (0.41 - 3.03) | 0.86 (0.39 - 1.93) | Not reported |
| Di Prinzio (2020) | Austrailia | Cohort | 1980-2001 | Psychiatric hospital admission for any severe mental illness prior to index pregnancy | State hospital registry and mental health information system | Severe mental illness | <20 - 35+ | 7508 | 239365 | Not reported | Not reported | OR | 1.2 (1.0 - 1.5) | 1.1(0.9 - 1.3) | Not reported |
| Dubey (2025) | USA | Cross sectional | 2017 | ICD-10 code | National hospital database | Bipolar disorder | Dec-55 | Not reported | Not reported | Not reported | Not reported | OR | 1.33 (1.00, 1.76) | 1.09 (0.82 - 1.45) | 1.05 (0.79 - 1.42) |
| Fabre (2021) | France | Cohort | 2015-2019 | ICD-10 code | National hospital discharge data | Schizophrenia | 15 - 35+ | 3108 | 3664353 | 59 | 106 | OR | 2.19 (1.63-2.94 | 2.17 (1.37 – 3.44) | Not reported |
| Mei-Dan (2015) | Canada | Cohort | 2003-2011 | ICD-9, ICD-10 | Provincial health dataset | Bipolar disorder | 14 - 50 | 1859 | 432358 | 11 | 2235 | OR | Not reported | 1.20 (0.66 - 2.18) | Not reported |
|  |  |  |  |  |  | Major depression disorder | 14 - 50 | 3724 | 432358 | 16 | 2235 | OR | Not reported | 0.90 (0.55 - 1.47) | Not reported |
| Christopher (2023) | USA | Cohort | 2003-2018 | ICD-10 codes | Electronic health record database | Mental health disorders | 18 - 44 | Not reported | Not reported | Not reported | Not reported | OR | 1.71(1.58 - 1.85) | Not reported | Not reported |

Appendix 3: Joanna Briggs Institute Quality Appraisal Checklists

Cohort studies

| **Study Author (Year)** | **1. Were the two groups similar and recruited from the same population** | **2. Were the exposures measured similarly to assign people to both exposed and unexposed groups?** | **3. Was the exposure measured in a valid and reliable way?** | **4.Were confounding factors identified?** | **5. Were strategies to deal with confounding factors stated?** | **6.Were the groups/participants free of the outcome at the start of the study (or at the moment of exposure)?** | **7. Were the outcomes measured in a valid and reliable way?** | **8. Was the follow up time reported and sufficient to be long enough for outcomes to occur?** | **9. Was follow up complete, and if not, were the reasons to loss to follow up described and explored?** | **10. Were strategies to address incomplete follow up utilized?** | **11. Was appropriate statistical analysis used?** | **Overall Score** | **Quality Rating** |
| --- | --- | --- | --- | --- | --- | --- | --- | --- | --- | --- | --- | --- | --- |
| Akobirshoev (2024) | Yes | Yes | Yes | Yes | Yes | Yes | Yes | Partially | Partially | Partially | Yes | 73 | High |
| Mitra (2021) | Yes | Yes | Yes | Yes | Yes | Yes | Yes | Partially | Partially | Partially | Yes | 73 | High |
| Gleason (2021) | Yes | Yes | Yes | Yes | Yes | Yes | Yes | Partially | Partially | Partially | Yes | 73 | High |
| Zhong (2018) | Yes | Yes | Yes | Yes | Yes | Yes | Yes | Partially | Partially | Partially | Yes | 73 | High |
| Brown (2022) | Yes | Yes | Yes | Yes | Yes | Yes | Yes | Partially | Partially | Partially | Yes | 73 | High |
| Mei-Dan (2015) | Yes | Yes | Yes | Yes | Yes | Yes | Yes | Partially | Partially | Partially | Yes | 73 | High |
| Di Prinzio (2020) | Yes | Yes | Yes | Yes | Yes | Yes | Yes | Partially | Partially | Partially | Yes | 73 | High |
| Fabre (2021) | Yes | Yes | Yes | Yes | Yes | Yes | Yes | Yes | Yes | Yes | Yes | 100 | High |
| White (2016) | Yes | Yes | Yes | Yes | Yes | Yes | Yes | Yes | Partially | Partially | Yes | 82 | High |
| Mitra (2020) | Yes | Yes | Yes | Yes | Yes | Yes | Yes | Partially | Partially | Partially | Yes | 73 | High |
| Akorbirshoev (2017) | Yes | Yes | Yes | Yes | Yes | Yes | Yes | Partially | Partially | Partially | Yes | 73 | High |
| Shin (2020) | Yes | Yes | Yes | Yes | Yes | Yes | Yes | Partially | Partially | Partially | Yes | 73 | High |
| Sundelin (2018) | Yes | Yes | Yes | Yes | Yes | Yes | Yes | Yes | Yes | Yes | Yes | 100 | High |
| Easter (2019) | Yes | Yes | Yes | Yes | Yes | Yes | Yes | Partially | Partially | Partially | Yes | 73 | High |

Cross-sectional studies:

| **Study Author (Year)** | **1. Were the criteria for inclusion in the sample clearly defined?** | **2. Were objective, standard criteria used for measurement of the condition?** | **3. Was the exposure measured in a valid and reliable way?** | **4. Were the outcomes measured in a valid and reliable way?** | **5. Were confounding factors identified?** | **6. Were strategies to deal with confounding factors stated?** | **7. Was appropriate statistical analysis used?** | **8.Were the study subjects and the setting described in detail?** | **Overall Score** | **Quality Rating** |
| --- | --- | --- | --- | --- | --- | --- | --- | --- | --- | --- |
| Akobirshoev (2021) | Yes | Yes | Partially | Partially | Yes | Yes | Yes | Yes | 75 | High |
| Dubey (2025) | Yes | Yes | Yes | Yes | Yes | Yes | Yes | Yes | 100 | High |

Appendix 4: List of Excluded Studies

| Author, year | Title | Reason for exclusion |
| --- | --- | --- |
| Accortt et al., 2022 | Association between diagnosed perinatal mood and anxiety disorders and adverse perinatal outcomes | No comparison |
| Acharya et al., 2015 | Preventing maternal deaths and overcoming challenges related to disability in pregnant women | Wrong article type |
| Adane et al., 2021 | Disparities in severe neonatal morbidity and mortality between Aboriginal and non-Aboriginal births in Western Australia: A decomposition analysis | No disability |
| Adane et al., 2021 | The impact of maternal prenatal mental health disorders on stillbirth and infant mortality: a systematic review and meta-analysis | Wrong article type |
| Adeoye et al.,2025 | Determinants of adverse perinatal outcomes in Ibadan, Nigeria. The influence of maternal lifestyle. | No disability |
| Admon et al., 2018 | Obstetric outcomes and delivery-related health care utilization and costs among pregnant women with multiple chronic conditions | No disability |
| Akobirshoev et al., 2024 | Severe maternal morbidity by disability status and type in the United States | Wrong outcomes |
| Akobirshoev et al., 2019 | Racial and ethnic disparities in birth outcomes and labor and delivery-related charges among women with intellectual and developmental disabilities | No comparison |
| Arechvo et al., 2023 | Incidence of stillbirth: effect of deprivation | No disability |
| Blackman et al., 2024 | Severe maternal morbidity and mental health hospitalizations or emergency department visits | Wrong outcomes |
| Brown et al., 2026 | Multiple maternal chronic conditions and risk of severe neonatal morbidity and mortality | No disability |
| Brown et al., 2026 | Multiple chronic conditions and risk of adverse maternal health outcomes: Population-based cohort study | No disability |
| Chan et al., 2025 | Adverse obstetric and neonatal outcomes associated with maternal schizophrenia-spectrum disorders and prenatal antipsychotic use: a meta-analysis of 37,214,330 pregnancy deliveries and propensity-score weighted population-based cohort study assessing confounder dependency of risk estimates | Wrong outcomes |
| Chuu et al., 2019 | Racial disparities in maternal and neonatal outcomes among pregnant women with bipolar disorder | No comparison |
| Dawson et al., 2022 | Social determinants and inequitable maternal and perinatal outcomes in Aotearoa New Zealand | Wrong outcomes |
| DeJong et al., 2018 | Pregnancy in patients with bipolar disorder: Maternal and neonatal outcomes | Wrong outcomes |
| Easter et al., 2021 | Obstetric near misses among women with serious mental illness: Data linkage cohort study | Wrong outcomes |
| Goldacre et al., 2017 | Women with intellectual disability are at a higher risk of adverse maternal and offspring outcomes | Wrong article type |
| Hilder et al., 2025 | Using linked data to assess maternal mental and behavioral disorders in pregnancy | No full article |
| Howland et al., 2016 | A population-based study of severe maternal morbidity in New York City, 2008-2012 | Wrong outcomes |
| Lo et al., 2025 | Pregnancy and postnatal outcomes for women with intellectual disability and their infants: A systematic review | Wrong article type |
| McKee et al., 2020 | Perinatal mood and anxiety disorders, serious mental illness and delivery-related health outcomes, United States, 2006-2015 | Wrong outcomes |
| Mitra et al., 2019 | Severe maternal morbidity among women with intellectual and developmental disabilities: A population-based study | Wrong outcomes |
| Mitra et al., 2016 | Fetal outcomes among US women with intellectual and developmental disabilities | Incomplete results |
| Mueller et al., 2019 | Pregnancy course, infant outcomes, rehospitalization, and mortality among women with intellectual disability. | Wrong outcomes |
| Nishat et al., 2022 | Continuity of primary care and prenatal care adequacy among women with disabilities in Ontario: a population-based cohort study. | Wrong outcomes |
| O'Mahen et al., 2024 | Obstetric and neonatal outcomes in pregnant women with and without a history of specialist mental health care: a national population-based cohort study using linked routinely collected data in England. | No full text |
| Parekh et al., 2025 | Risk of Cardiovascular Events During Delivery Hospitalization Among Pregnant Patients with Mental Health Conditions. | Wrong outcomes |
| Razaz et al., 2019 | Perinatal outcomes in women with multiple sclerosis: a population-based cohort study in Sweden. | Wrong outcomes |
| Schiff et al., 2021 | Pregnancy outcomes among visually impaired women in Washington State, 1987–2014. | Wrong outcomes |
| Schiff et al., 2017 | Pregnancy outcomes among deaf women in Washington State, 1987-2014 | Wrong outcomes |
| Schneiderman et al., 2017 | Maternal and neonatal outcomes of pregnancies in women with Addison's disease: a population‐based cohort study on 7.7 million births. | No disability |
| Signore et al., 2021 | The intersection of disability and pregnancy: risks for maternal morbidity and mortality. | Wrong article type |
| Singh et al., 2021 | Trends and racial/ethnic, socioeconomic, and geographic disparities in maternal mortality from indirect obstetric causes in the United States, 1999-2017. | No disability |
| Ward et al., 2025 | Maternal ethnic group, socioeconomic status, and neonatal and child mortality: a nationwide cohort study in England and Wales. | No disability |

Appendix 5: Funnel Plot for Maternal Mortality Meta-analysis

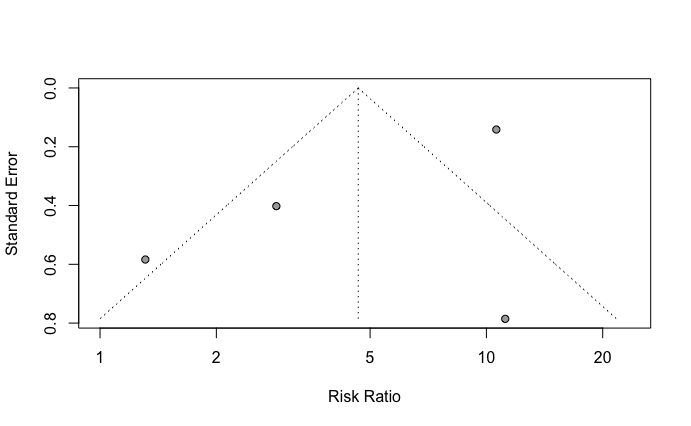

Appendix 6: Funnel Plot for Neonatal Mortality Meta-analysis

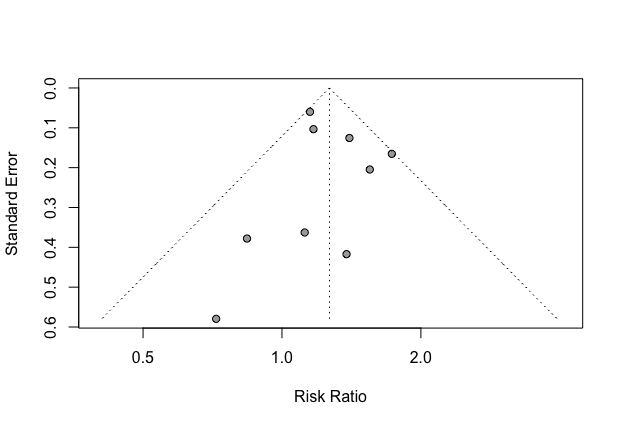

Appendix 7: Funnel Plot for Infant Mortality Meta-analysis

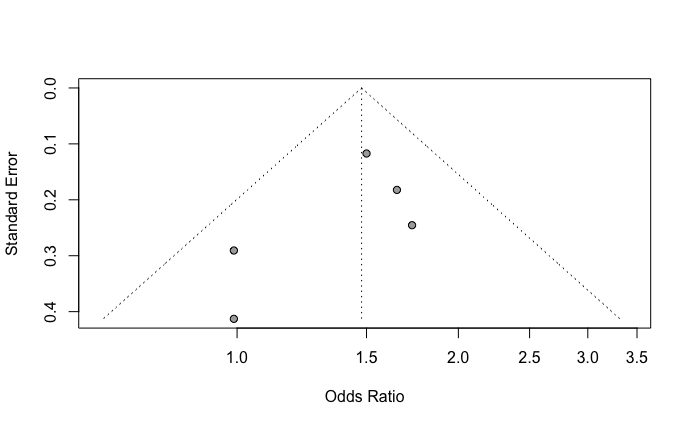

Appendix 8: Funnel Plot for Stillbirth Meta-analysis

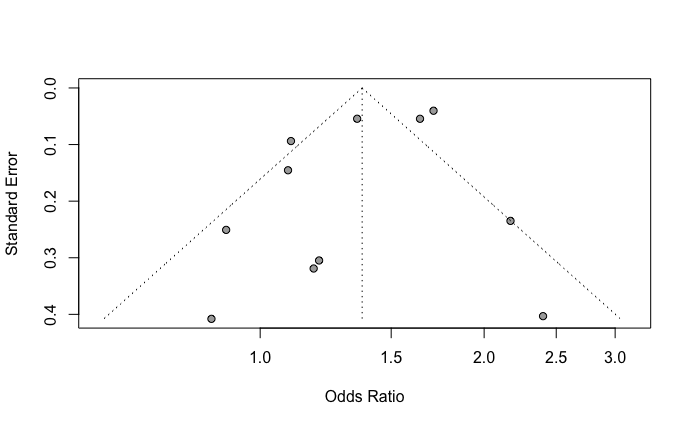
